# Assessing the Stability of Photon-Counting CT: Insights from a Two-Year Longitudinal Study

**DOI:** 10.1101/2024.06.05.24308046

**Authors:** Leening P. Liu, Pouyan Pasyar, Fang Liu, Quy Cao, Olivia F. Sandvold, Martin V. Rybertt, Pooyan Sahbaee, Russell T. Shinohara, Harold I. Litt, Peter B. Noël

## Abstract

**Objective:** Among the advancements in computed tomography (CT) technology, photon-counting computed tomography (PCCT) stands out as a significant innovation, providing superior spectral imaging capabilities while simultaneously reducing radiation exposure. Its long-term stability is important for clinical care, especially longitudinal studies, but is currently unknown. This study sets out to comprehensively analyze the long-term stability of a first-generation clinical PCCT scanner.

**Methods:** Over a two-year period, from November 2021 to November 2023, we conducted weekly identical experiments utilizing the same multi-energy CT protocol. Throughout this period, notable software and hardware modifications were meticulously recorded. Various tissue-mimicking inserts were scanned weekly to rigorously assess the stability of Hounsfield Units (HU) and image noise in Virtual Monochromatic Images (VMIs) and iodine density maps.

**Results:** Spectral results consistently demonstrated the quantitative stability of PCCT. VMIs exhibited stable HU values, such as variation in relative error for VMI 70 keV measuring 0.11% and 0.30% for single-source and dual-source modes, respectively. Similarly, noise levels remained stable with slight fluctuations linked to software changes for VMI 40 and 70 keV that corresponded to changes of 8 and 1 HU, respectively. Furthermore, iodine density quantification maintained stability and showed significant improvement with software and hardware changes, especially in dual-source mode with nominal errors decreasing from 1.44 to 0.03 mg/mL. Conclusion This study provides the first long-term reproducibility assessment of quantitative PCCT imaging, highlighting its potential for the clinical arena.

**Key Points:** **Question:** Photon-counting CT (PCCT) provides critical spectral imaging for improved diagnostic accuracy, but its long-term quantitative stability over time is still unknown.

**Findings:** The clinical PCCT system demonstrated stable Hounsfield Units (HU) and image noise over two years, ensuring reliable quantitative imaging and improving diagnostic accuracy.

**Clinical Relevance:** This study showcased the exceptional value of PCCT in diagnostic radiology, particularly for its application in longitudinal studies.

## Introduction

Follow-up and surveillance using computed tomography (CT) are essential for several medical conditions, including oncology, infectious diseases, and cardiothoracic disorders. In oncology, CT is pivotal for detecting and monitoring recurrences or metastases, such as in lung cancer post-treatment^1^ and colorectal cancer^2^. In infectious diseases, CT scans are utilized in the follow-up of tuberculosis (TB) to assess treatment response, especially in pulmonary TB, and to monitor for complications like cavitation and fibrosis^3^. For chronic diseases, CT is employed to monitor disease progression, detect complications, and assess treatment response in chronic obstructive pulmonary disease^4^. In cardiovascular diseases, surveillance via CT is crucial for monitoring aortic aneurysm size and growth, which is essential for timely surgical intervention^5^. Quantitative imaging through CT has become increasingly important for these tasks, especially in longitudinal studies, with the introduction of new CT systems, such as photon-counting CT (PCCT)^6–9^.

PCCT represents a significant advancement in diagnostic imaging technology, offering a more detailed view of various anatomical and pathological structures. Unlike traditional CT scanners, PCCT employs photon-counting detectors that can measure the energy of individual X-ray photons. This innovation allows for enhanced spectral imaging, enabling differentiation between different tissues and materials^10,11^. PCCT provides high-resolution images with improved contrast^12–15^, reduces need for additional scans^16–18^, and minimizes radiation exposure for patients^19–21^ compared to both traditional CT and dual-energy CT scanners. Since the introduction of the first PCCT system in Europe in spring 2021^22^ and its FDA approval in October 2021, clinical impacts of the technology have been successfully demonstrated. One significant advantage lies in enhanced quantitative capabilities, primarily attributed to spectral imaging, which enables the reduction of influences caused from both patient and scanner, such as beam hardening artifacts. This advancement has been exemplified through virtual monoenergetic images (VMI) and iodine density maps, especially with clinical prototype systems^23–25^. Additionally these spectral maps are always available with PCCT without specific protocol selection, which facilitates translation of these maps for clinical applications. However, in light of ongoing advancements, it is essential to assess the long-term precision and robustness of PCCT’s improved quantitative capabilities.

Though photon-counting detectors are a promising and innovative technology, they signify a significant leap beyond incremental changes in CT technology. Thoroughly assessing their long-term stability and usability is essential. Additionally, when compared to conventional CT detector technology, photon-counting detectors introduce new sensitivities, such as cooling requirements and system temperature management, that may affect quantitative accuracy over time. Therefore, this study aims to conduct a comprehensive analysis of the long-term stability of a first-generation clinical PCCT scanner. Our findings demonstrate the consistent quantitative stability of a clinical PCCT system over an extended period, offering important insights into its long-term performance and reliability.

## Materials and Methods

### Phantom

A phantom (Multi-energy CT phantom, Gammex, Sun Nuclear) was utilized to evaluate the quantitative stability of PCCT over time (**Figure 1**). The phantom was scanned in its full form consisting of an inner phantom with a diameter of 20 cm and an oval extension ring measuring 30 x 40 cm. It contains bores for material-specific and tissue-mimicking inserts. Specifically, the inner phantom included the following inserts: adipose, brain, blood 40, blood 70, blood 100, blood + 2 mg/mL iodine, blood + 4 mg/mL iodine, calcium 50 mg/mL, iodine 2 mg/mL, iodine 5 mg/mL, and iodine 10 mg/mL. The extension ring included calcium 100 mg/mL, calcium 300 mg/mL, 2 mm iodine core, 5 mm iodine core, and 10 mm iodine core inserts. To compare the measured values, ground truth values were calculated for different photon energies (40, 70, 100, 190 keV) using phantom manufacturer provided elemental composition and physical densities:

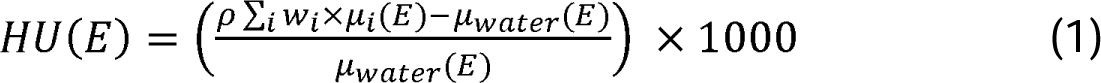

where *E* is the photon energy, *ρ* is the physical density, *w_i_* is the mass fraction for a specific element *i*, *µ_i_* is the mass attenuation coefficient of element *i* at energy *E*, and *µ_water_* is the mass attenuation coefficient of water at energy *E*.

**Figure 1.**
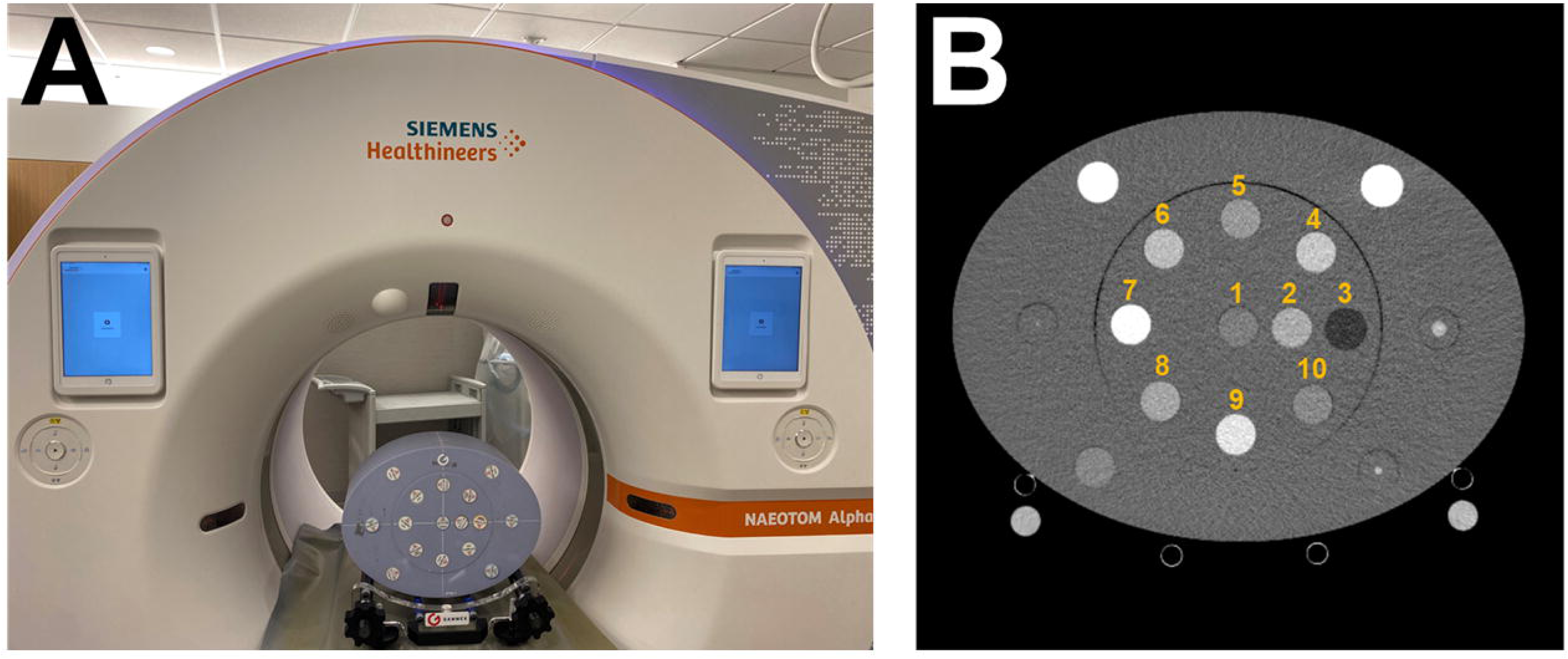
Experimental set up. A phantom with an extension ring (**A**) was scanned with different tissue and material specific inserts. Regions of interests were then placed on specific inserts (**B**) for analysis.

#### Image acquisition

The phantom was scanned on a dual-source PCCT (NAEOTOM Alpha, Siemens Healthineers) approximately every week between November 23, 2021 and November 22, 2023 excluding holidays, vacations, and service events, averaging 1.3 weeks between scans. Any software updates and significant hardware updates were additionally recorded to investigate quantitative deviations associated with these changes. Of note, notable software updates occurred at weeks 8, 35, and 69, while notable hardware changes were implemented at weeks 8, 19 and 80. Brief descriptions of software updates were provided. These descriptions highlighted an improvement in cross-scatter correction at week 8 and 35 but no other specific improvements besides general image quality improvements. Scans were performed with the phantom in the isocenter of the scanner in both single source and dual source modes at a tube voltage of 120 kVp and a volumetric CT dose index (CTDI_vol_) of 10 mGy. Scans in each source mode were repeated a total of three times to account for statistical variation. For each scan, VMI 40, 70, 100, and 190 keV as well as iodine density maps were reconstructed with a field of view of 450 mm, a slice thickness of 3 mm, and a reconstruction filter of Qr48. To assess temporal variation in noise with and without denoising, spectral results were reconstructed at two different levels of quantum iterative reconstruction (QIR): 0 and 2. Other acquisition and reconstruction parameters can be found in **Table 1**.

**Table 1.** Acquisition and reconstruction parameters.

| Scanner model | NAEOTOM Alpha |
| --- | --- |
| Tube voltage | 120 kVp |
| Source modes | Single-source, dual-source |
| Rotation time | 0.25 s |
| Pitch | 1 |
| Collimation | 144 x 0.4 mm |
| CTDI<sub>vol</sub> | 10 mGy |
| Slice thickness | 3 mm |
| QIR | 0, 2 |
| Reconstruction filter | Qr48 |
| Reconstructed field of view | 450 mm |
| Matrix size | 512 x 512 |
| Pixel spacing (in x and y) | 0.88 mm |

#### Image analysis

To examine temporal stability of photon-counting CT, ten consecutive central slices from each scan were analyzed. Regions of interest (ROI) were first automatically placed on a dual-source VMI 70 keV with in-house software and then were copied to other spectral results for the same week. For each ROI, mean and standard deviation (noise) were measured. The mean value and mean noise were then calculated across the three repeated scans (30 slices total/week). Additionally, the percent error relative to the ground truth and absolute error was calculated for each insert and VMI at each time point. For iodine density, the nominal and absolute error relative to the ground truth was calculated. Temporal variation in relative error was then visualized in a scatter plot separately for single and dual source modes with a QIR level of 0. Noise was similarly represented in a scatter plot over time with noise from spectral results with QIR level 0 and 2. In figures that display results over time, significant software updates and hardware changes are indicated by a gray bar. To quantitatively characterize variation, the mean and standard deviation of the measured values over time for different inserts and parameters was calculated across the entire duration of the study as well as between hardware and software changes.

#### Statistical analysis

To identify changes in quantification, the Pruned Exact Linear Time (PELT) algorithm^26^, a change point search algorithm, was utilized for each unique combination of spectral result, iterative reconstruction, source type, and insert. This algorithm identified times where the statistical properties changed, i.e. changes in mean, and was applied to both metrics, relative error (nominal or percent) and noise. These change points were then tallied up across inserts for different combinations of parameters to determine major and minor breakpoints for each spectral result and metric.

## Results

### VMI temporal stability

VMIs from 40 to 190 keV exhibited quantitative stability over time. Specifically, VMI 70 keV values in single-source mode demonstrated no significant changes over time for any of the inserts (**Figure 2**). The overall relative error in single-source mode averaged to 0.86% ± 0.54%, which is equivalent to 9.6 ± 6.6 HU across different inserts, while the variation of VMI 70 keV in single-source mode averaged 0.11% across the inserts. Mean absolute errors over the two-year period ranged from 2 to 9 HU for different inserts with the exception of calcium 50 mg/mL, which demonstrated an absolute error of 20 and 28 HU for single-source and dual-source modes, respectively (**Table 2**). Variation of the mean absolute error, on the other hand, ranged from 1 to 4 HU, highlighting PCCT’s quantitative stability. In comparison, VMI 70 keV values in dual-source mode experienced an overall variation of 0.30%. The difference in variation between VMI 70 keV in single-source and dual-source modes can be attributed to identified change points in dual-source mode at week 8 for a majority of the inserts (8/10, **Figure 2**). It resulted in decrease in VMI 70 keV values from a relative error of 1.42% to 0.06% across all inserts that may have been a consequence of both hardware and software changes to improve cross-scatter correction. In addition, change points were also identified at week 35 for brain and blood 100 inserts, and at week 69 for blood + 4 mg/mL iodine and iodine 10 mg/mL inserts. These change points varied for VMIs for different energies (**Figure S1-3**). In particular, the software update at week 35 also caused change points for VMI 40 (8/10), 100 (6/10), and 190 keV (10/10) and 40 keV (6/10) for single-source and dual-source modes, respectively. Similar to VMI 70 keV, relative error in dual-source mode was altered in VMI 40 (10/10), and 190 keV (8/10). A minor effect was also displayed for VMI 40 keV in single-source mode at week 8 (2/10). Other change points were identified at timepoints with no software or hardware changes but largely only occurred for a single insert.

**Figure 2.**
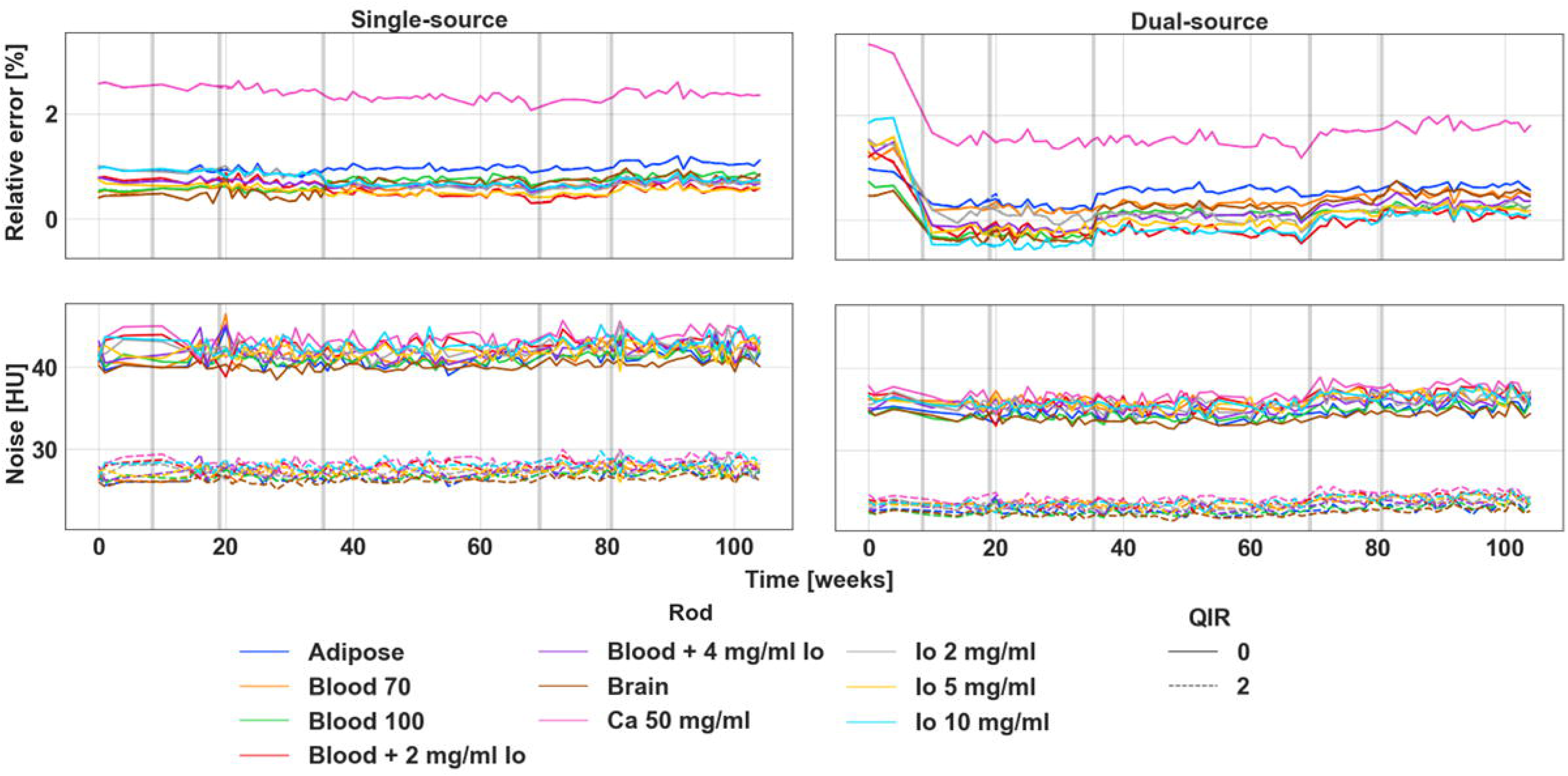
Stability of VMI 70 keV relative error and noise across time for single-source and dual-source modes. Single-source quantification was largely stable with no change points identified while dual-source quantification improved significantly with change points at week 8. Additionally, minor change points were determined at weeks 35 and 69 that only affected 2/10 inserts each. Noise, both in single-source and dual-source modes, remained stable with an identified change point at week 69 for dual-source mode that corresponded to a decrease in noise by approximately 1 HU. Gray bars in the figure indicate significant software (weeks 8, 35, 69) and hardware updates (weeks 8, 19, 80). VMI: virtual monoenergetic images, HU: Hounsfield Units, QIR: quantum iterative reconstruction.

**Table 2.**
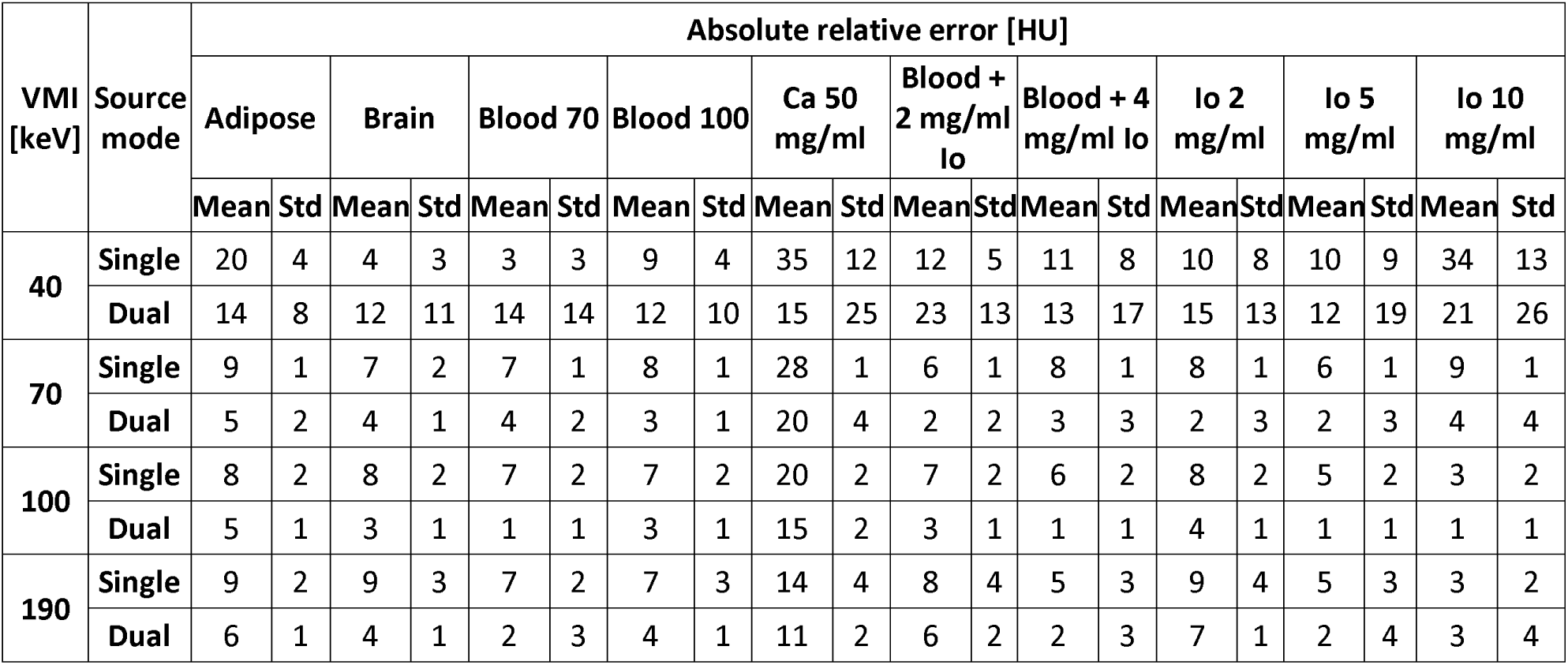
Mean absolute nominal error for different VMIs over a two-year period.

| VMI<br>[keV] | Source<br>mode | Absolute relative error [HU] |  |  |  |  |  |  |  |  |  |  |  |  |  |  |  |  |  |  |  |
| --- | --- | --- | --- | --- | --- | --- | --- | --- | --- | --- | --- | --- | --- | --- | --- | --- | --- | --- | --- | --- | --- |
|  |  | Adipose |  | Brain |  | Blood 70 |  | Blood 100 |  | Ca 50<br>mg/ml |  | Blood +<br>2 mg/ml<br>lo |  | Blood + 4<br>mg/ml lo |  | lo 2<br>mg/ml |  | lo 5<br>mg/ml |  | lo 10<br>mg/ml |  |
|  |  | Mean | Std | Mean | Std | Mean | Std | Mean | Std | Mean | Std | Mean | Std | Mean | Std | Mean | Std | Mean | Std | Mean | Std |
| 40 | Single | 20 | 4 | 4 | 3 | 3 | 3 | 9 | 4 | 35 | 12 | 12 | 5 | 11 | 8 | 10 | 8 | 10 | 9 | 34 | 13 |
|  | Dual | 14 | 8 | 12 | 11 | 14 | 14 | 12 | 10 | 15 | 25 | 23 | 13 | 13 | 17 | 15 | 13 | 12 | 19 | 21 | 26 |
| 70 | Single | 9 | 1 | 7 | 2 | 7 | 1 | 8 | 1 | 28 | 1 | 6 | 1 | 8 | 1 | 8 | 1 | 6 | 1 | 9 | 1 |
|  | Dual | 5 | 2 | 4 | 1 | 4 | 2 | 3 | 1 | 20 | 4 | 2 | 2 | 3 | 3 | 2 | 3 | 2 | 3 | 4 | 4 |
| 100 | Single | 8 | 2 | 8 | 2 | 7 | 2 | 7 | 2 | 20 | 2 | 7 | 2 | 6 | 2 | 8 | 2 | 5 | 2 | 3 | 2 |
|  | Dual | 5 | 1 | 3 | 1 | 1 | 1 | 3 | 1 | 15 | 2 | 3 | 1 | 1 | 1 | 4 | 1 | 1 | 1 | 1 | 1 |
| 190 | Single | 9 | 2 | 9 | 3 | 7 | 2 | 7 | 3 | 14 | 4 | 8 | 4 | 5 | 3 | 9 | 4 | 5 | 3 | 3 | 2 |
|  | Dual | 6 | 1 | 4 | 1 | 2 | 3 | 4 | 1 | 11 | 2 | 6 | 2 | 2 | 3 | 7 | 1 | 2 | 4 | 3 | 4 |

### VMI noise

VMI noise was mostly invariant across 104 weeks, with small changes in noise attributed to software changes. For VMI 70 keV, noise was stable at 42 ± 1 and 27 ± 1 HU in single-source mode and 35.5 ± 0.9 and 23.4 ± 0.6 HU in dual-source mode for QIR 0 and 2, respectively (**Figure 2**). While an increase in noise occurred after the hardware change at week 35, it was temporary and did not persist over time. However, a change point was detected at week 68 for dual-source mode in 7/10 and 3/10 inserts, corresponding to average change in noise across all inserts of 1.3 and 0.9 HU for QIR 0 and 2, respectively. With this small change, overall, the variation as measured by standard deviation was 0.9 and 0.6 HU for both single-source and dual-source modes for QIR 0 and 2, respectively. Comparatively, VMI 40 keV contained change points at weeks 8, 35, and 69 (**Figure S1**). In particular, changes at week 35 affected all inserts for all combinations of source type and QIR level with decreases of 7, 8, 7, and 9 HU for single-source + QIR 0, single-source + QIR 2, dual-source + QIR 0, and dual-source + QIR 2, respectively. Quantitative changes from week 8 and 69, on the other hand, paralleled that of VMI 70 keV noise with a magnitude of 2 HU. Additionally, no major change points were identified for VMI 100 and 190 keV for any combination of source types and QIR levels.

### Iodine density

The quantification of iodine density demonstrated significant improvement and stability with both software updates and hardware changes, particularly in dual-source mode (**Figure 3**). In single-source mode, only one change point was identified at week 35 where the average nominal error fell from 0.31 to -0.04 mg/mL (**Figure 3**). On the other hand, two separate change points were identified in dual-source mode (**Figure 3**). The biggest change occurred at week 8, falling from an average nominal error of 1.44 to -0.27 mg/mL, and was observed for all five analyzed iodine inserts. The second change point at week 69 was only ascertained for 3/5 iodine inserts with a change in nominal error of 0.21 mg/mL. Ultimately, these changes evened out the difference in nominal error between source modes with nominal errors of -0.06 and 0.03 mg/mL for single-source and dual-source modes, respectively. Overall, absolute error across the two years ranged from 0.10 ± 0.06 to 0.38 ± 0.21 mg/mL for different iodine-containing inserts (**Table 3**). Beside the change points, quantitative values demonstrated stability through the mean standard deviation of nominal differences ranging from 0.04 to 0.12 mg/mL for the different periods between software and/or hardware changes. Similar stability was seen in single-source mode, with standard deviations ranging from 0.02 to 0.07 mg/mL. Iodine density noise also remained relatively stable but with a significant change at week 35, changing from 0.63 to 1.33 and 0.48 to 0.85 mg/mL for QIR 0 and 2, respectively (**Figure 4**).

**Figure 3.**
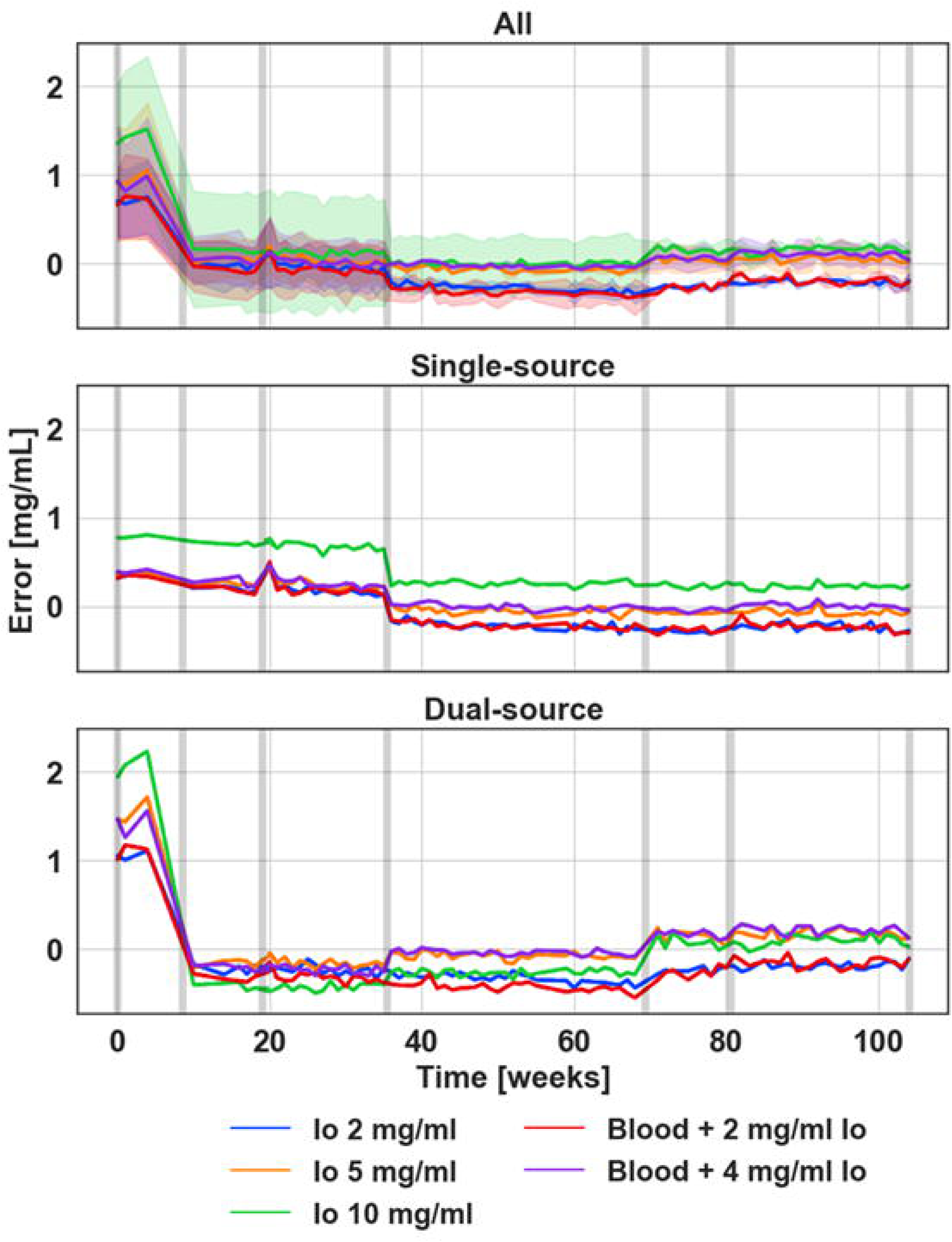
Temporal variations in relative nominal error in iodine density quantification. Combined single-source and dual-source data, where shaded regions correspond to the standard deviation across different combinations of source types and IR levels, demonstrated reduction in the variance between iodine density between source type and, ultimately, convergence of quantification as a result of software and hardware updates. Iodine density in single-source mode remained mostly invariant with the exception of one change point at week 35 while iodine density in dual-source mode exhibited significant changes that improved its accuracy. Gray bars in the figure indicate significant software (weeks 8, 35, 69) and hardware updates (weeks 8, 19, 80).

**Table 3.** Mean absolute nominal error for iodine maps over a two-year period.

| Source mode | Absolute relative error [mg/mL] |  |  |  |  |  |  |  |  |  |
| --- | --- | --- | --- | --- | --- | --- | --- | --- | --- | --- |
|  | Blood + 2<br>mg/ml lo |  | Blood + 4<br>mg/ml lo |  | lo 2 mg/ml |  | lo 5 mg/ml |  | lo 10 mg/ml |  |
|  | Mean | Std | Mean | Std | Mean | Std | Mean | Std | Mean | Std |
| **Single** | 0.22 | 0.06 | 0.10 | 0.06 | 0.22 | 0.06 | 0.12 | 0.10 | 0.38 | 0.21 |
| **Dual** | 0.34 | 0.19 | 0.20 | 0.19 | 0.28 | 0.17 | 0.18 | 0.27 | 0.31 | 0.37 |

**Figure 4.**
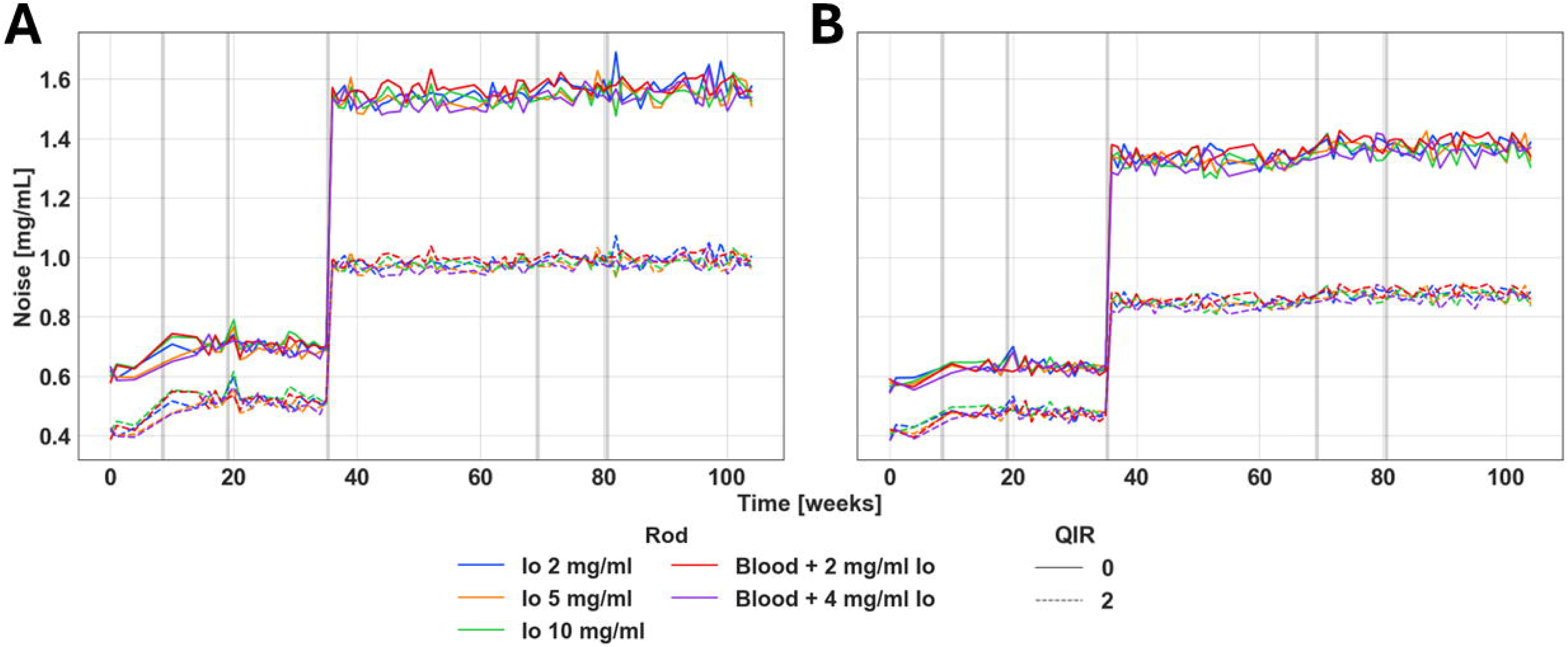
Iodine density noise over time in single-source and dual-source modes. Iodine noise in single-source (**A**) and dual-source (**B**) modes displayed temporal stability between software and hardware changes. A notable increase in noise in both source types was visible at week 35. Gray bars in the figure indicate significant software (weeks 8, 35, 69) and hardware updates (weeks 8, 19, 80).

## Discussion

In this paper, we present the first study demonstrating long-term reproducibility of quantitative PCCT imaging. We illustrate that this clinical system, the first of its kind, has significantly improved over time and has become a robust and stable imaging tool. In addition to its enhanced stability, our study highlights several key findings that further validate the effectiveness of PCCT for quantitative imaging applications. Notably, the system consistently performs soundly with both single- and dual-source acquisition protocols. While previous technologies, such as energy-integrating detectors, offer similar overall consistency, they are limited compared to PCCT, which provides advantages such as the removal of patient- and system-dependent artifacts via spectral, ultra-high resolution, and low-dose imaging.

PCCT is on its way to becoming a diagnostic imaging tool in the coming years. The current body of literature suggests that various vendors are preparing distinct technical implementations for clinical deployment^24,27,28^. Although the longitudinal performance of these forthcoming systems remains untested, the PCCT assessed in this work has demonstrated quantitative stability over time crucial for longitudinal studies and other applications. It is also anticipated that these future systems will share similar advantages, thus enabling the consistent assessment of disease progression for patients with cancer and chronic diseases. Furthermore, as PCCT becomes more widely available, its combination of longitudinal stability and high-resolution imaging opens up possibilities in radiomics. Radiomics involves converting digital medical images into mineable high-dimensional data, driven by the idea that these images contain information about underlying pathophysiology, which can be uncovered through quantitative image analyses^29^. Studies conducted by other investigators have highlighted the opportunities of merging radiomics with PCCT for a single imaging timepoint. These findings have shown that PCCT achieves greater estimation accuracy for morphological radiomics features than conventional CT systems, emphasizing the potential of this technology for improved quantitative imaging^30,31^. While challenges with respect to protocol and parameter harmonization for radiomics remain even with PCCT, opportunities to improve radiomics, particularly in oncological follow-up and surveillance, are enhanced with PCCT. Future studies to evaluate long-term radiomics stability using patient-based lifelike phantoms^32^ will be necessary.

This study has several limitations: (i) We only evaluated a single protocol designed for abdominal imaging. Further studies will be necessary to assess the long-term performance of technically demanding protocols, such as those used in cardiac imaging. (ii) We assessed only one PCCT system, for which we observed high reproducibility over time. In a follow-up study, we plan to evaluate inter-scanner stability to provide essential technology insight for planning multicenter clinical studies. (iii) We focused our evaluation on VMIs and iodine density maps. Other spectral results were not included in the study, but it is known that the performance of individual spectral results and maps is interconnected. (iv) The timeframe of this study also imposes a limitation since we acquired images for only about two years, while the lifetime of such an imaging tool is much longer. We are planning to continue this study and collect additional data to illustrate effects that may only become apparent over time. In summary, while this study provides valuable insights into the performance of a specific imaging protocol and a particular PCCT system, it is imperative to interpret the results within the context of these limitations.

PCCT marks a significant advancement in medical imaging, delivering superior image quality, reduced radiation exposure, and a wide range of clinical applications. This study indicates that the technology is quantitative stable over time, which is critical to its adoption as a tool in diagnostic imaging.

## Data Availability

All data produced in the present study are available upon reasonable request to the authors.

## Abbreviations

CT: computed tomography
TB: tuberculosis
PCCT: photon-counting CT
VMI: virtual monoenergetic images
HU: Hounsfield units
QIR: quantum iterative reconstruction
ROI: region of interest
PELT: pruned exact linear time

