## Supplementary material for "Assessing the Stability of Photon-Counting CT: Insights from a Two-Year Longitudinal Study": Figure S

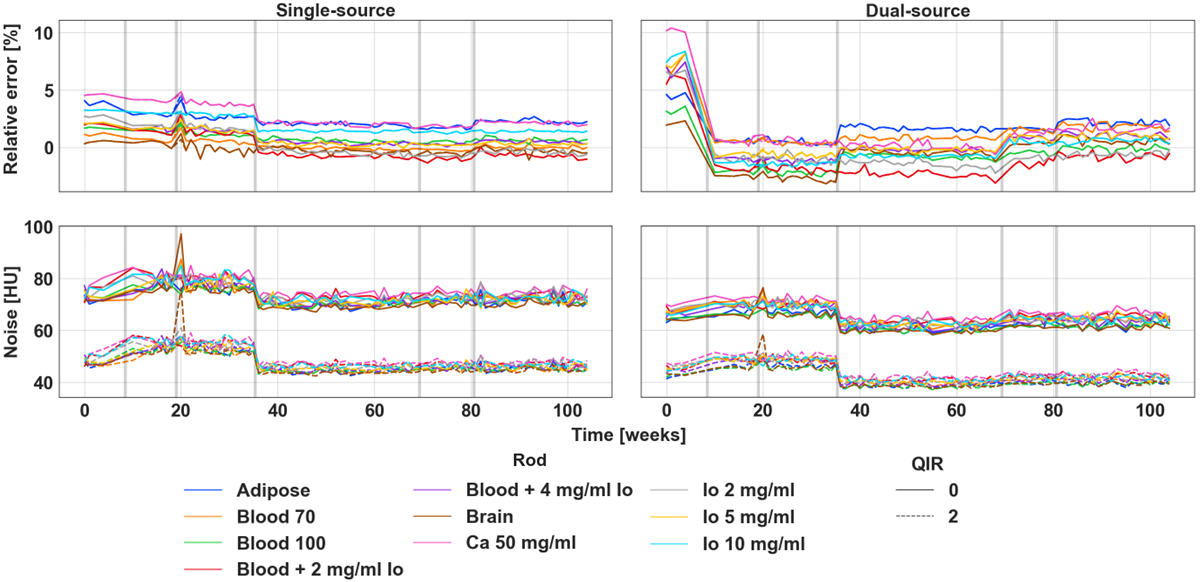

**Figure S1. Stability of VMI 40 keV relative error and noise across time for single-source and dual-source modes.** Gray bars in the figure indicate significant software (weeks 8, 35, 69) and hardware updates (weeks 8, 19, 80).

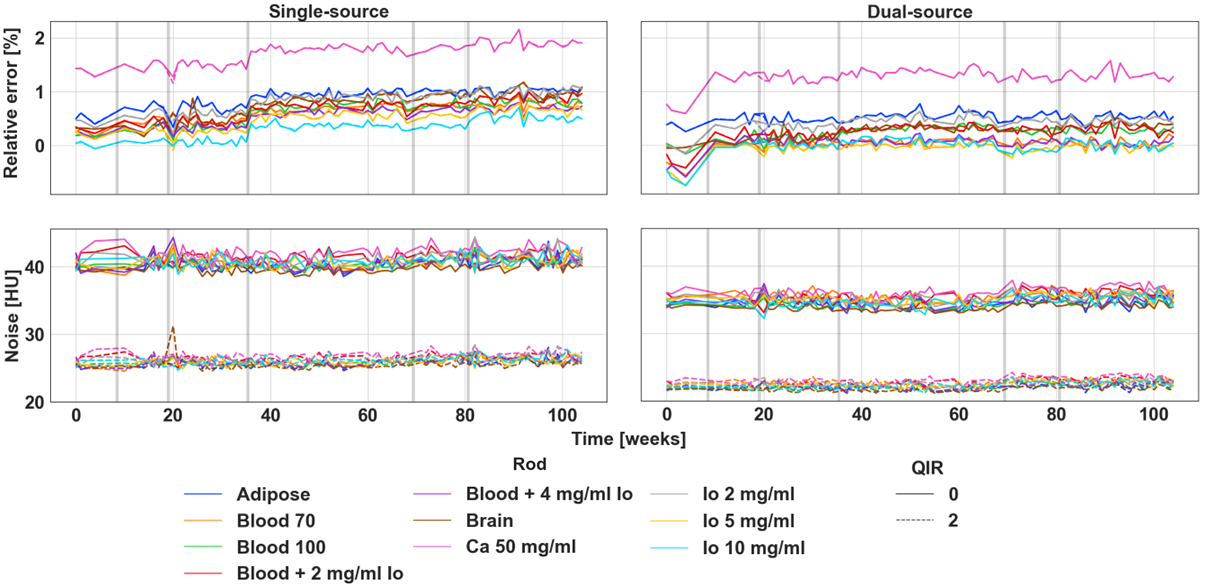

**Figure S2. Stability of VMI 100 keV relative error and noise across time for single-source and dual-source modes.** Gray bars in the figure indicate significant software (weeks 8, 35, 69) and hardware updates (weeks 8, 19, 80).

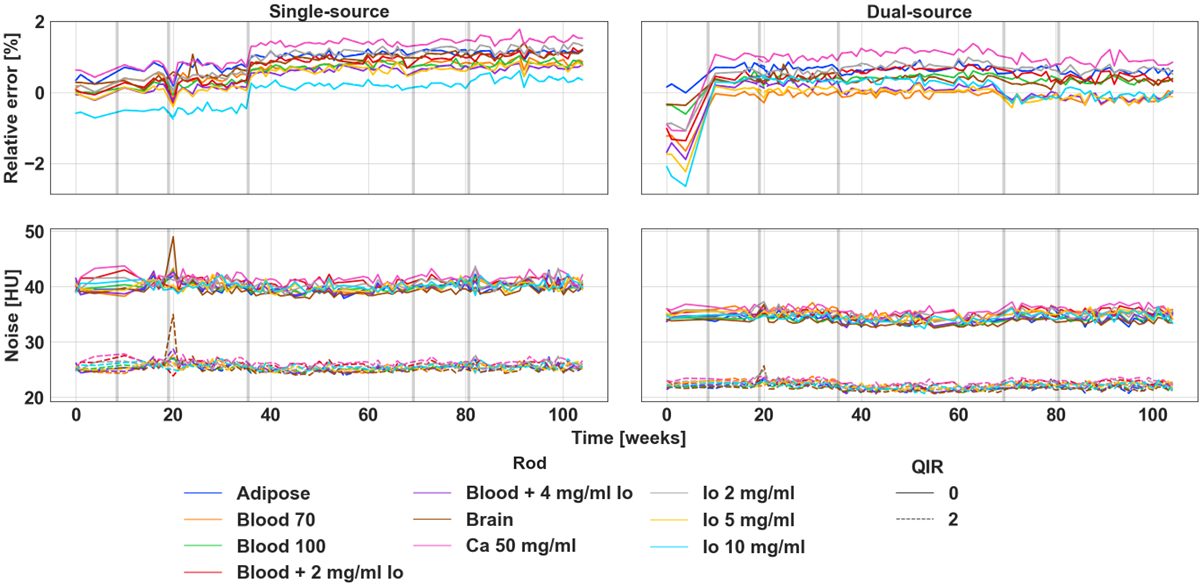

**Figure S3. Stability of VMI 190 keV relative error and noise across time for single-source and dual-source modes.** Gray bars in the figure indicate significant software (weeks 8, 35, 69) and hardware updates (weeks 8, 19, 80).
